# Protocol for a scoping review of literature on digital sexual, reproductive, and gender health care for LGBTQI+ youth

**DOI:** 10.1101/2023.08.25.23294615

**Authors:** Julie McLeod, Paul Flowers, Jo Gibbs, Claudia S. Estcourt, Jennifer MacDonald

## Abstract

**Introduction:** Lesbian, gay, bisexual, trans, queer, questioning, and other sexual and gender minority (LGBTQI+) youth are diverse populations who experience poor sexual health outcomes (e.g., high rates of sexually transmitted infections (STIs) and human immunodeficiency virus (HIV), unplanned pregnancy, and sexual violence) and multiple barriers to sexual and reproductive health care (SRHC) and gender health care (GHC). In high-income, developed countries, barriers include confidentiality concerns; stigma and discrimination; and health care providers’ lack of specific training. Digital SRHC and GHC have the potential to overcome key barriers for LGBTQI+ youth by offering anonymous and independent access to care. However, the literature on digital SRHC and GHC for LGBTQI+ youth is fragmented, often focussing on one sub-population at a time, despite shared barriers. The extent and nature of recent literature regarding digital SRHC and GHC for LGBTQI+ youth is unclear, as is acceptability of, and barriers/facilitators to, LGBTQI+ youth engaging with digital SRHC and GHC.

**Objective:** To identify, describe, and evaluate the methodological quality of, the existing literature on digital SRHC and GHC for LGBTQI+ youth in high-income, developed countries, synthesise study findings, and make recommendations for future research.

**Inclusion criteria:** Research studies from 2018 onward in published and grey literature on any aspect of digital (e.g., websites, mobile applications) SRHC and GHC (e.g., online information, support and advice, and clinical care for STIs and HIV, fertility, sexual violence, sexual wellbeing, and gender expression and transition) for LGBTQI+ youth (aged 10-35 years) in high-income, developed countries.

**Method:** This study will follow the Joanna-Briggs Institute (JBI) methodology for scoping reviews. The databases to be searched include APA PsycInfo (ProQuest); APA PsycArticles (ProQuest); CINAHL Complete (EBSCO); MEDLINE (EBSCO); ERIC (EBSCO); British Education Index (EBSCO); Education Database (ProQuest); Computer Science Database (ProQuest); and Web of Science. Grey literature will be identified using Google Scholar. Studies will be screened against and selected for inclusion in line with the eligibility criteria. Key data from included studies will be extracted to a structured spreadsheet, adapted from the JBI extraction tool, then synthesised qualitatively using the JBI meta-aggregative approach for a systematic narrative account, accompanied by tables as appropriate.

## Introduction

### The problem

Lesbian, gay, bisexual, trans, queer, questioning, intersex and other sexual orientation and gender diverse and minority (LGBTQI+) (Stonewall, n.d.; YoungScot, 2022) youth (broadly aged 10-35, as ‘youth’ is categorised in various age ranges across research, ranging from 10 to 34 years: e.g., Decker et al., 2021; Step et al., 2022) are heterogenous and marginalised populations facing to two key issues regarding sexual and reproductive health (SRH) and gender health (GH): 1) poor SRH and GH outcomes; and 2) barriers to sexual and reproductive health care (SRHC) and gender health care (GHC). See Tables 1 and 2 for an overview of abbreviations and key terms.

**Table 1.** Acronyms used throughout this protocol and their definitions.

| **Acronym** | **Definition** |
| --- | --- |
| GBMSM | Gay, bisexual, and other men who have sex with men |
| GH | Gender health |
| GHC | Gender health care |
| HCP | Health care provider |
| HIV | Human immunodeficiency virus |
| LGBTQI+ | Lesbian, gay, bisexual, transgender, queer, questioning, intersex, and other sexual orientation and gender diverse and minority populations |
| PrEP | Pre-exposure prophylaxis |
| SRH | Sexual and reproductive health |
| SRHC | Sexual and reproductive health care |
| STI | Sexually transmitted infection (e.g., chlamydia, gonorrhoea, syphilis) |
| UN | United Nations |
| WHO | World Health Organisation |

**Table 2.**
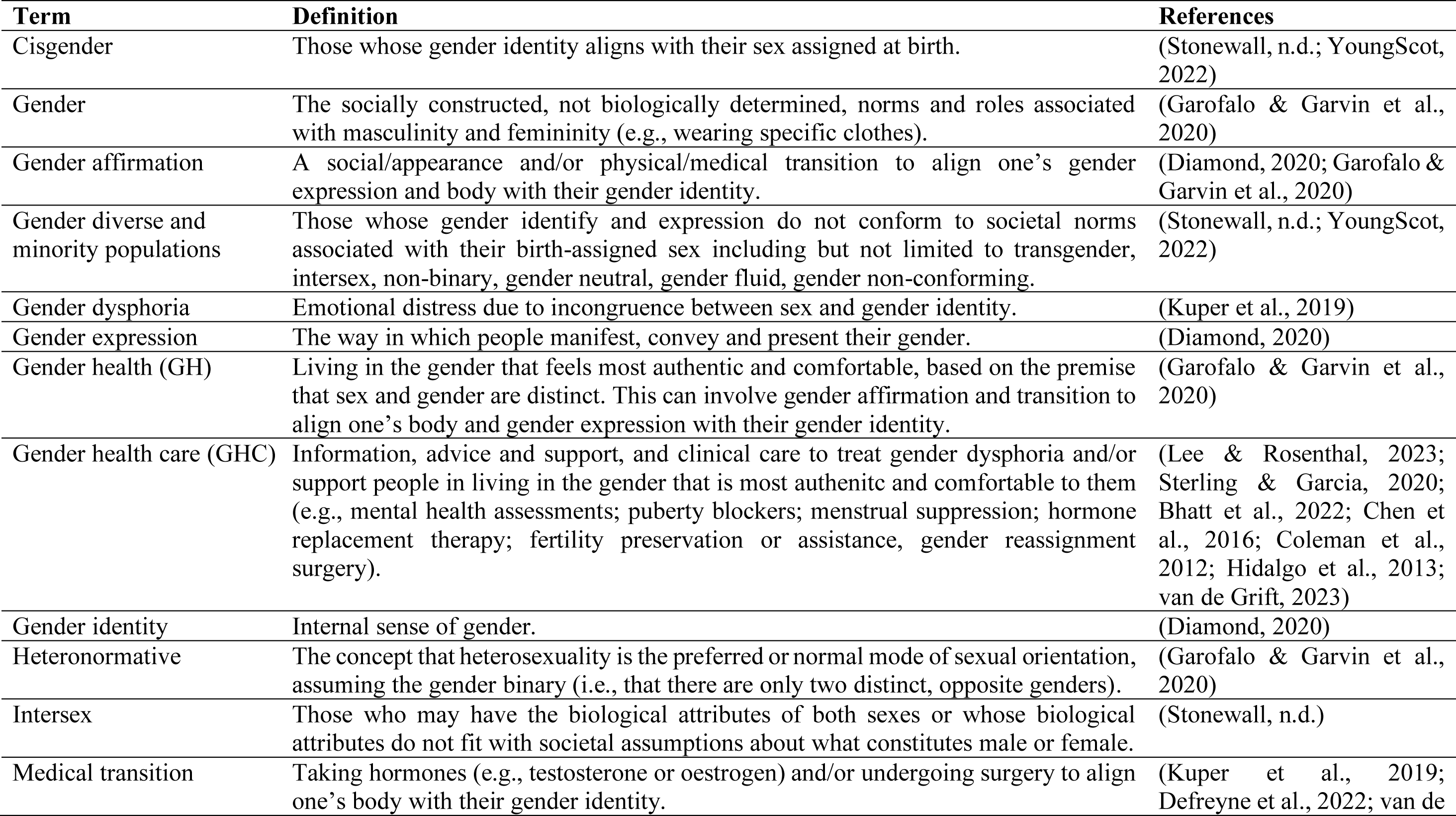
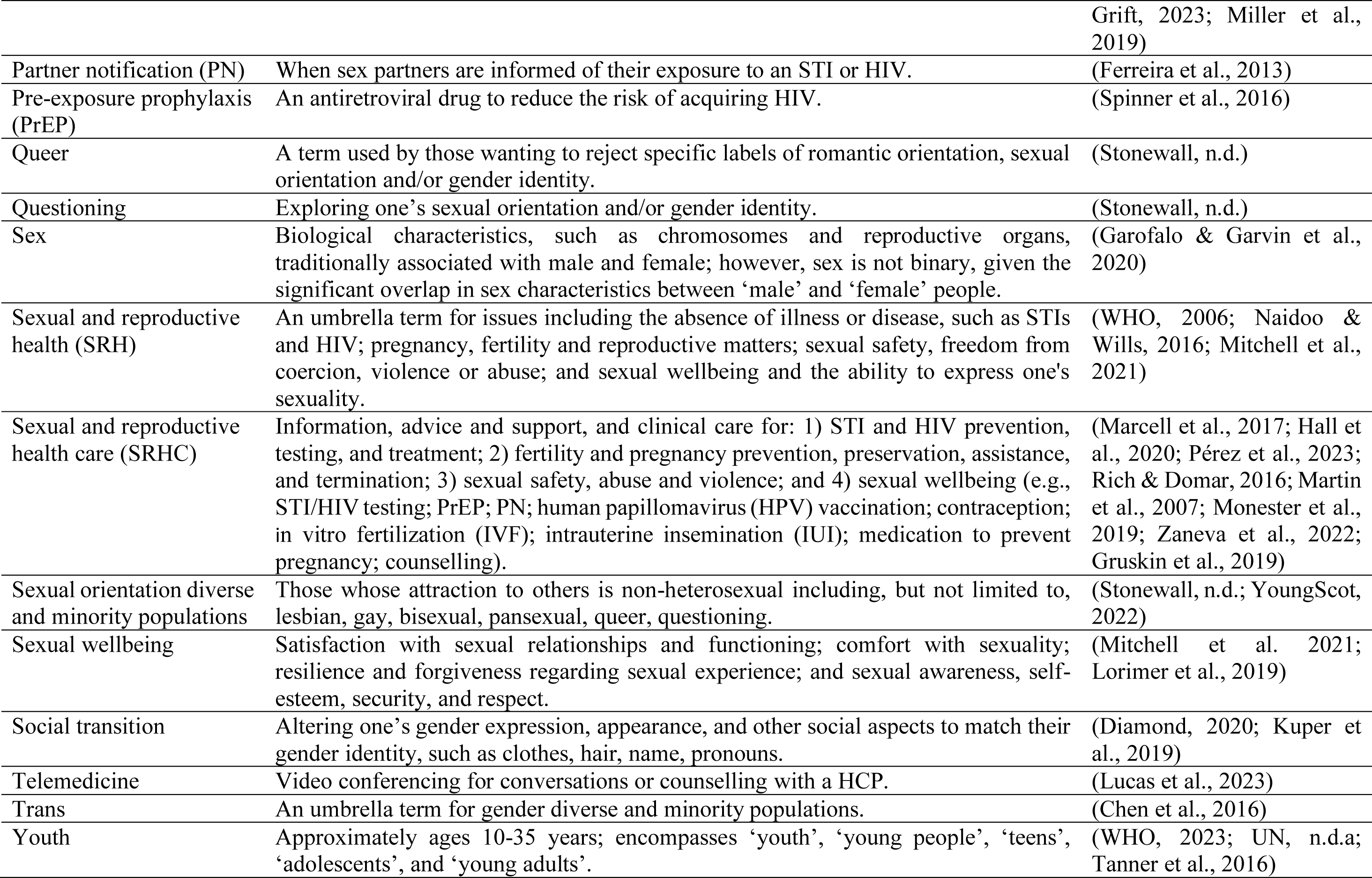
Glossary of terms used within this protocol, their definitions, and references.

Firstly, compared to their heterosexual, cisgender and adult counterparts, LGBTQI+ youth are at disproportionately high risk for acquiring sexually transmitted infections (STIs) and human immunodeficiency virus (HIV), particularly young gay, bisexual and other men who have sex with men (GBMSM) and trans women (Quinn et al., 2021; Reisner et al., 2019; UKHSA, 2022; Day et al., 2021; Epps et al., 2023). Additionally, LGBTQI+ youth disproportionately experience early and unplanned pregnancy, and sexual abuse and violence, particularly young bisexual women and trans youth (Wood et al., 2016; Ybarra et al., 2022; Quinn et al., 2021; Reisner et al., 2019; UKHSA, 2022; Day et al., 2021; Epps et al., 2023; Atteberry-Ash et al., 2020; Semprevivo et al., 2021; Saewyc, 2014; Tornello et al., 2014; Ela & Budnick, 2017; Eisenberg et al., 2020). Moreover, compared with heterosexual youth, LGBTQI+ youth are more likely to be sexually active, have earlier sexual debut (before age 13) and have four or more sexual partners, and are less likely use condoms (Wood et al., 2016; Arrington-Sanders et al., 2016). Alongside poor SRH outcomes, LGBTQI+ youth can experience gender dysphoria (emotional distress due to incongruence between gender and sex) and related anxiety and depression, and are at high risk of self-harm and suicidal behaviours (Tan et al., 2022; Sood et al., 2021). LGBTQI+ youth have been found to experience higher levels of anxiety, depression, and discomfort with their gender identity based on societal norms and expectations than LGBTQI+ adults (Jackman, 2018).

Secondly, LGBTQI+ youth experience barriers to engaging with SRHC and GHC and often have low help-seeking behaviour (Phillips et al., 2019; Sharma et al., 2019; Boydell et al., 2017; Hibbert et al., 2020; Newcomb et al., 2020; Wood et al., 2016). Barriers include confidentiality concerns; perception and experience of discrimination, marginalisation and stigma; health care providers’ (HCPs) lack of knowledge and training about their needs; and heteronormative assumptions of their gender and sexuality (Hudson-Sharp & Metcalf, 2016; Fisher et al., 2018; Boydell et al., 2017; Lefkowitz & Mannell, 2017; Mkhize & Maharaj, 2020; Logie et al., 2019; Quinn et al., 2019; Knight et al., 2014). For GHC specifically, a key barrier is also long wait times of months to years (Puckett et al., 2018; Scheim et al., 2021; Wright et al., 2021). Long wait times can lead to LGBTQI+ youth self-administering hormones acquired from the internet, friends, family, and strangers, without medical consultation, to undergo physical transition to align their body with their gender identity (Mepham et al., 2014; Rotondi et al., 2013; Kennedy et al., 2021). Self-administering hormones is problematic as it is associated with less awareness of side effects than obtaining hormones from a physician and severe medical complications, such as cardiovascular disease or cancer (Mepham et al., 2014; Metastasio et al., 2018).

It is important to consider SRH and GH together, as research indicates that GH is a critical issue relating to SRH. Gender dysphoria can negatively impact LGBTQI+ people’s sexual behaviour and expression, due to discomfort with their genitals, such as never allowing sex partners to touch their genitals or breasts (Gil-Llario et al., 2021). Further, receiving GHC can positively impact all types of sexual experiences and activities among trans people (Bungener et al., 2020) Additionally, trans people who have received GHC are more likely than those who have not to have sex and to do so more often (Nikkelen & Kreules, 2018). Gender affirmation is also associated with increased STI testing and awareness of the HIV prevention drug, pre-exposure prophylaxis (PrEP) (Andrzejewski et al., 2021). However, little is known about the relationship between SRH and GH among LGBTQI+ youth, as much of the research has focussed on adult populations (e.g., Gil-Lario et al., 2021; Bungener et al., 2020; Nikkelen & Kreules, 2018; Andrzejewski et al., 2021).

It is also essential to consider the intersectionality of gender and sexual orientation with other socio-economic demographics, such as ethnicity, occupation and education, as these intersecting demographics can be associated with increased risk of poor outcomes and barriers to engaging with health care (O’Neill et al., 2014; Ng, 2016). For example, in the United Kingdom (UK) and United States (US), Black populations are disproportionately affected by STIs and HIV (UKHSA, 2022; Hightow-Weidman et al., 2012; Jones, 2020). Additionally, intersectional stigma experienced by young LGBTQI+ people of colour in the US and Canada can impact identity disclosure to HCPs and decisions about uptake of PrEP (Quinn et al., 2019; Shanshan, 2014).

### A potential solution

Digital SRHC and GHC, of which there has been a rapid expansion in recent years (Norman et al., 2022; Ong et al., 2022; Tucker et al., 2022; Robinson & Johnston, 2018; Yousaf & Currie, 2021; Lunt et al., 2021), offers care that is private, self-managed, and convenient, overcoming key barriers to accessing health care for LGBTQI+ youth (Magee et al., 2012; Mustanski et al., 2015; Stephenson et al., 2020; Refugio et al., 2019; Bauermeister et al., 2022; DeHaan et al., 2013). An initial literature search indicates that digital SRHC and GHC includes: 1) *information* delivered online on websites (e.g., World Health Organisation [WHO], 2022; Magee et al., 2012) or through online education programmes (e.g., Mustanski et al., 2015; Lacombe-Duncan et al., 2021); 2) *advice and support* online which may involve use of telecommunication such as video calls, live chats, email, or text services with charity or community-based organisation staff or peers (e.g., Radix et al., 2022; Skeen & Cain, 2022); and 3) *clinical care* provided online, such as STI/HIV self-sampling kits and test results (e.g., Bauermeister et al., 2015; Estcourt et al., 2017; Hottes et al., 2012; Ling et al., 2010; Brugha et al., 2011); PrEP (ePrEP; Kincaid et al., 2021); partner notification (PN) and testing options (e.g., Golden et al., 2022; Flowers et al., 2021; Mimiaga et al., 2008); and consultations (eConsult) and telemedicine (e.g., Sequeira et al., 2021; Lucas et al., 2023).

Digital SRHC and GHC have been found to be preferable to in-person services among LGBTQI+ youth (Cherenack et al., 2016; Flanders et al., 2017; Gilbert et al., 2013) and LGBTQI+ youth often turn to the internet for SRH information to compensate for lack of formal education or a trusted person to ask (DeHaan et al., 2013; Mitchell et al., 2014; Bradford et al., 2019). Thus, it is important to understand the barriers and facilitators to digital SRHC and GHC among LGBTQI+ youth in order to determine how to best to deliver SRHC and GHC for LGBTQI+ youth. However, the extent of existing digital SRHC and GHC *for LGBTQI+ youth* is unclear, as the research does not often focus specifically on LGBTQI+ youth, but rather general populations (e.g., Bailey et al., 2015; Guse et al., 2012; Maloney et al., 2020; Ortiz-Martínez & Ríos-González, 2017). Additionally, digital SRHC and GHC interventions for LGBTQI+ youth typically target specific sub-populations such as young GBMSM (e.g., Lelutiu-Weinberger et al., 2015) or transgender youth (e.g., Sequeira et al., 2020). As a result, the literature is fragmented, despite research indicating that LGBTQI+ youth face similar barriers to engaging with digital SRHC and GHC, for example: privacy concerns; lack of privacy at the time of engagement; the cognitive burden of finding, understanding, and interpreting information alone; low SRH literacy; and inequalities in access and skills to use and understand digital technology (Ybarra et al., 2020; Cherenack et al., 2016; Ventuneac et al., 2020; Epps et al., 2023; Robinson et al., 2020). Therefore, a review is needed to outline the scope of digital SRHC and GHC for LGBTQI+ youth, and their acceptability of, and barriers and facilitators to, their use. This scoping review seeks to fill this gap and identify knowledge to form the foundation of understanding how to optimise SRHC and GHC for LGBTQI+ youth. An important consideration is that barriers to SRHC and GHC (digital and non-digital) differ considerably between countries, depending on access to the internet and digital technology (Robinson et al., 2020), as well as infrastructure and social welfare/protections for access to health care (Germain et al., 2015; Gottlieb et al., 2014). Thus, this scoping review will focus on high-income, developed countries, such as the UK, Canada and Australia, for maximum generalisability of the findings. A systematic scoping review approach (e.g., Peters et al., 2015) was adopted as this was the most appropriate method to enable a mapping of the literature to examine the type and range of research activity and to identify gaps in the literature regarding digital SRHC and GHC for LGBTQI+ youth (Peters et al., 2020; Arksey & O’Malley, 2005).

A preliminary search of MEDLINE, the Cochrane Database of Systematic Reviews, Joanna-Briggs Institute (JBI) Evidence Synthesis, and British Medical Journal (BMJ) Open using the words “LGBT youth sexual health”, “LGBT youth STI”, and “LGBT youth STD” was conducted (31.01.2023) and no published or ongoing systematic or scoping reviews on the topic were identified. However, three similar reviews were identified during the literature search (Gilbey et al., 2020; Cui et al., 2022; and Stoehr et al., 2022). See Appendix A for an overview of how this current study differs from these three studies.

### Review objective

The objective of this scoping review is to identify, describe, and evaluate the methodological quality of, the existing literature on digital SRHC and GHC for LGBTQI+ youth in high-income, developed countries, synthesise study findings, and make recommendations for future research.

### Review questions (RQs)

RQ1: What dimensions of digital SRHC and GHC for LGBTQI+ youth in high-income, developed countries have received attention in the literature and who are the target populations?
RQ2: What are the characteristics of LGBTQI+ youth in research regarding digital SRHC and GHC and accessing and using digital SRHC and GHC in high-income, developed countries?
RQ3: What is the acceptability of digital SRHC and GHC in high-income, developed countries for LGBTQI+ youth?
RQ4: What are the barriers and facilitators to LGBTQI+ youth accessing and using digital SRHC and GHC in high-income, developed countries?
RQ5: How is LGBTQI+ ‘youth’ defined in digital SRHC and GHC research from high-income, developed countries?
RQ6: How, if at all, have theory or frameworks been used in research into digital SRHC and GHC for LGBTQI+ youth in high-income, developed countries?

## Eligibility criteria

The proposed scoping review will be conducted in accordance with the JBI methodology for scoping reviews, using the participant, concept, context (PCC) framework (Peters et al., 2020).

### Participants

#### Inclusion

The populations of focus in this scoping review are LGBTQI+ youth.

##### LGBTQI

Studies with all LGBTQI+ gender and sexual orientation diverse and minority identifying participants will be included in this scoping review. This will be inclusive of all terms to describe sexual minorities and gender diverse populations, such as non-binary, gender non-conforming, gender fluid, and gender neutral (Stonewall, n.d.).

##### Youth

Here, ‘youth’ encompasses ‘young people’, ‘young adults’ and ‘adolescents’ and covers an age range of 10 to 35 years. There is no universal definition or standardised operationalisation of ‘youth’ or its synonyms used in research. The World health Organisation (WHO) and United Nations (UN) define ‘youth’ as 15-24 years, ‘young people’ as 10-24 years and ‘adolescents’ as 10-19 years (WHO, 2023; UN, n.d.a). However, research into SRHC and GHC for LGBTQI+ youth employs a wider age range between 10 to 35 years (e.g., 10-20, Decker et al., 2021; 13-29, Wood et al., 2016; 16-24, Magee et al., 2012; 16-29, Flanders et al., 2017; 18-26, McRee et al., 2015; 15-34, Step et al., 2022). Additionally, ‘youth’ is categorised in various age ranges across countries’ legal and policy frameworks and research, ranging from 10 to 29 years (Perovic, 2016). Moreover, those of gender and sexuality minority populations have been found to have sex earlier than heterosexual and cisgender counterparts (Wood et al., 2016), including 10 years of age (Arrington-Sanders et al., 2016). Therefore, the age range in this scoping review is deliberately broad to understand how ‘youth’ is operationalised in digital SRHC and GHC literature.

#### Exclusion

Research focussing on non-LGBTQI+ populations will be excluded. Studies will also be excluded if data for LGBTQI+ participants are not/cannot be disaggregated from non-LGBTQI+ participant data. Studies with child or adult populations (aged below 10 or above 35 years), or where data from participants within the age range of 10-35 years cannot be disaggregated from child/adult populations, will also be excluded.

### Concept

#### Inclusion

The overarching concept of interest for this scoping review is digital SRHC and GHC.

##### Digital

‘Digital’ encompasses all internet-based and digitally mediated SRHC and GHC including, but not limited to, websites; web applications (apps); mobile apps; text messaging or short messaging service (SMS); email; and video conferencing (e.g., Holloway et al., 2014; Mustanski et al., 2015). Where studies report on both digital and non-digital SRHC and GHC and it is possible to disaggregate the digital from the non-digital data, these studies will be included. Where services offer hybrid in-person and digital options, these will be included.

##### SRHC

For this scoping review, SRHC will incorporate the provision of, or engagement with, information, advice and support, and clinical care for LGBTQI+ youth regarding SRH (e.g., STIs/HIVs, fertility/pregnancy, and sexual violence, prevention, testing, treatment and management, and sexual wellbeing). SRHC will be inclusive of studies of services and interventions. Services refer to digital SRHC designed for LGBTQI+ youth for help-seeking, such as information, telecommunication, or clinical care (e.g., online STI/HIV self-sampling kits; PN; PrEP; contraception; counselling; forums, social media groups). Interventions refer to digital strategies that have been developed to change a specific SRH related behaviour(s) or outcome(s) for LGBTQI+ youth populations, such as education programmes (Mustanski et al., 2015), social marketing campaigns (e.g., Hickson et al., 2015), or signposting to local services (e.g., West et al., 2015) (e.g., to increase condom use, sexual health related knowledge, uptake of STI self-sampling kits) including offering a digital version of a service that is typically delivered in person (e.g., Brady et al., 2015).

##### GHC

For this scoping review, GHC will incorporate the provision of, or engagement with information, advice and support, and clinical care for LGBTQI+ youth regarding GH (e.g., gender identity, expression, affirmation, and social or physical transition). This will be inclusive of research into services and interventions. Services refer to digital GHC designed for LGBTQI+ youth for help-seeking, such as information, telecommunication, or clinical care (e.g., electronic consultation (e-consult); counselling; and access to hormone replacement therapy (HRT) (Radix et al., 2022). Interventions refer to digital strategies that have been developed to change a specific GH related behaviour or outcome for LGBTQI+ youth, such as education programmes, (e.g., to increase knowledge and understanding or decision making about GHC; Salvetti et al., 2022), including digital versions of services typically delivered in person (e.g., Shipherd et al., 2016; Blosnich et al., 2019).

#### Exclusion

Non-digital SRHC and GHC delivered in-person or by phone will be excluded. Where studies report on both digital and non-digital SRHC and GHC and it is not possible to disaggregate the digital from the non-digital data, these studies will be excluded. Additionally, where studies focus on the recruitment of participants (e.g., digital methods of recruitment to digital or non-digital SRHC or GHC interventions, or digital/non-digital recruitment to digital SRHC or GHC interventions), these studies will be excluded.

Studies reporting on digital care for mental health (e.g., anxiety, depression) for LGBTQI+ youth will also be excluded. While anxiety and depression are key mental health issues experienced by LGBTQI+ youth, particularly trans youth, this is distinct from gender health, relating specifically to gender identity, expression, affirmation and transition. Gender affirmation and receiving GHC have been found to improve the mental health and wellbeing of LGBTQI+ youth, reduce anxiety, depression, and suicidal ideation and behaviour (Russell et al., 2018; Sorbara et al., 2020; Tordoff et al., 2022).

### Context

#### Inclusion

Studies from high-income and developed economy countries, as defined by the UN (2023a), will be included: Australia; Austria; Belgium; Canada; Croatia; Cyprus; Czech Republic; Denmark; Estonia; Finland; France; Germany; Greece; Hungary; Iceland; Ireland; Italy; Japan; Latvia; Lithuania; Luxembourg; Malta; Netherlands; New Zealand; Norway; Poland; Portugal; Slovakia; Slovenia; Spain; Sweden; Switzerland; United Kingdom; and United States.

While there is a move away from the terms ‘developed’ and ‘developing’ (Jimba et al., 2019), these are still the terms currently used by the UN as of 2023 (UN, 2023a, 2023b). These inclusion criteria have been applied as this scoping review is part of a wider PhD programme of formative research aiming to provide theoretically informed and evidence-based guidance for how to optimise digital SRHC and GHC for LGBTQI+ youth in the UK. Therefore, these context inclusion criteria are necessary to ensure the findings from included studies will be maximally generalisable and applicable across countries similar to the UK, with regards to access to the internet and digital technology and the provision of social policies which support access to health care (Robinson et al., 2020; Quaglio et al., 2016; UN, 2021, n.d.b; Germain et al., 2015; Gottlieb et al., 2014; OECDStatistics, 2023, 2019; International Labour Organisation [ILO], 2021, 2022; Iyer, 2021; Kanchoochat, 2019; European Commission, n.d.; Azzopardi-Muscat et al., 2017; Konkov, 2017; Robles & Vargas, 2012; Arbulo et al., 2015).

#### Exclusion

Studies will be excluded if they were conducted in low-and-middle-income counties or countries with economies in transition and developing economies (UN, 2019). This is to ensure the studies included will be maximally generalisable and applicable across countries with similar economic and social policies to the UK, as this scoping review is part of a wider PhD programme seeking to optimise digital SRHC and GHC for LGBTQI+ youth.

### Types of Sources

#### Inclusion

This scoping review will consider qualitative, quantitative and mixed methods studies that are classified as original research, with primary data collection. Here, quantitative research encompasses experimental and quasi-experimental study designs including randomised controlled trials, non-randomised controlled trials, before and after studies, and interrupted time-series studies. In addition, analytical observational studies including prospective and retrospective cohort studies, case-control studies, longitudinal, and analytical cross-sectional studies will be considered for inclusion. This review will also consider descriptive observational study designs including case series, individual case reports and descriptive cross-sectional studies for inclusion. Qualitative studies will include research designs such as phenomenology, grounded theory, ethnography, qualitative description, and action research. Studies using theory-based implementation and behavioural science for intervention development and evaluation will also be considered for inclusion. Pilot and feasibility studies will also be included.

#### Exclusion

Reviews, conference abstracts, posters, registered reports, blogs, guidelines, text and opinion papers, letters, editorials, commentaries, protocols, preprints, and doctoral and master’s theses will be excluded.

### Quality appraisal

A quality appraisal will be conducted on included papers that report on the acceptability of, or barriers or facilitators to, digital SRHC and GHC among LGBTQI+ youth using the Mixed Methods Appraisal Tool version 2018 (Hong et al., 2018). A quality appraisal is not common practice for scoping reviews, due to the nature of scoping reviews to map literature, rather than analyse data and make inferences or draw conclusions using the outcomes of studies (Peters et al., 2020). However, in this scoping review, we intend to synthesise data and comment on the levels of acceptability and the barriers and facilitators to engaging with digital SRHC and GHC, as it is not expected that sufficient literature will be identified to justify a follow up systematic review. Therefore, a quality appraisal will be performed to assess how well included studies were conducted (Higgins et al, 2019) and will be considered when summarising the evidence base and drawing conclusions.

## Methods

The proposed scoping review will be conducted in accordance with the JBI methodology for scoping reviews (Peters et al., 2020).

### Search strategy

The search strategy will aim to locate both published studies and grey literature. The PCC framework will be used to structure the searches. However, only the ‘Participants’ and ‘Concept’ inclusion criteria will be used to structure the search. ‘Context’ will not be included in the search strategy but will be used to screen papers for inclusion and will be included in data extraction. The search strategy was developed in three steps:

Step 1: A preliminary search of MEDLINE, the Cochrane Database of Systematic Reviews, JBI Evidence Synthesis, and BMJ Open using the words “LGBT youth sexual health”, “LGBT youth STI”, and “LGBT youth STD” was conducted (31.01.2023) to identify articles on the topic. An analysis of the text words contained in the title and abstract of retrieved papers, and of the index terms used to describe the articles was conducted to develop a full search strategy, adaptable for the relevant databases/information sources. This was first developed for MEDLINE (see Appendix B). A second search using all identified key words and index terms will be undertaken across all included databases: APA PsycInfo (ProQuest); APA PsycArticles (ProQuest); CINAHL Complete (EBSCO); MEDLINE (EBSCO); ERIC (EBSCO); British Education Index (EBSCO); Education Database (ProQuest); Computer Science Database (ProQuest); and Web of Science. Five papers identified from the literature search as meeting the inclusion criteria will be used to ensure the search terms return relevant literature (Ventuneac et al., 2020; Ybarra et al., 2020; Sequeira et al., 2021; McRee et al., 2018; Bauermeister et al., 2022).

Step 2: To identify grey literature, a search using select words and phrases from the search terms – at least one from each search string (see Appendix B) – will be undertaken on Google Scholar.

Step 3: The reference list of studies identified as relevant to this review (i.e., those included) will be screened for additional sources.

#### Inclusion

Studies published in English and from 2018 onwards will be included, as data older than this may not be relevant in the rapidly expanding and changing field of digital SRHC and GHC (Norman et al., 2022). Furthermore, in 2018, the Scottish Government published a strategy for enhancing and transforming health and social services through delivery of health care digitally (Robinson & Johnston, 2018; Yousaf & Currie, 2021). Additionally, LGBTQI+ rights have been subject to much discussion and change over the past five years and may impact provision of and access to SRHC and GHC (McDermott et al., 2021). The date restriction of 2018 onwards will return a collection of the most up-to-date and studies, conducted in the previous five years.

#### Exclusion

Studies published in any language other than English will be excluded, due to lack of resources to support translation to other languages. Studies published before 2018 will also be excluded to ensure only the most up-to-date information is included in this scoping review, relevant to current global cultural and social climates (McDermott et al., 2021).

### Study/Source of Evidence selection

Following the searches within each database, all identified citations will be exported to excel and uploaded to Rayyan (Ouzzani et al., 2016) and duplicates removed. Rayyan was selected as this software has been found to have the best sensitivity for accurately identifying duplicates compared to Ovid multifile search, EndNote desktop, Mendeley, Zotero, and Covidence in a search of MEDLINE, Embase, Cochrane Central Register of Controlled Trials, and PsycINFO databases (McKeown & Mir, 2021).

The titles and abstracts of deduplicated studies will be screened (100% by the first author, JML, and 1% by a second reviewer, Ron O’Kane (RO)) for assessment against the eligibility criteria outlined above and categorised into one of three categories: ‘included’, where it is clear from the title and abstract that the study meets the inclusion criteria; ‘excluded’, where it is clear from the title and abstract that the study does not meet the inclusion criteria; and ‘maybe’, where it is unclear from the title and abstract if the study meets the inclusion criteria. The titles and abstracts screened by RO will be ordered from most to least relevant, using the ‘Compute Ratings’ function within Rayyan (Ouzzani et al., 2016), to prioritise the most relevant studies.

Following title and abstract screening, the full text of ‘included’ and ‘maybe’ studies will be assessed in detail against the inclusion criteria (100% by JML and 10% by RO). Reasons for exclusion of studies at each stage will be recorded and reported in the scoping review and categorised by participants (e.g., not LGBTQI+ youth), concept (e.g., not digital SRHC or GHC), context (e.g., not a high-income, developed country), study type (e.g., not original research or intervention development/evaluation), and publication type (e.g., not a journal article). Any uncertainty that arises at each stage of the selection process will be resolved through discussion with independent experienced reviewers (authors, JMD, PF, CSE, and JG). The results of the search and the study inclusion process will be reported in full in the final scoping review and presented in a Preferred Reporting Items for Systematic Reviews and Meta-Analyses extension for Scoping Review (PRISMA-ScR) flow diagram (Tricco et al., 2018).

### Data Extraction

Data from final included papers will be extracted to excel (by JML) using a data extraction tool adapted (by JML, see Appendix C) from the JBI Manual for Evidence charting table for data extraction synthesis (Peters et al., 2020). Data extraction will be guided by the PCC framework and will include specific details about the study methods, participants, concept, context, and key findings relevant to the review questions. If appropriate, authors of papers will be contacted to request missing or additional data, where required. The extraction tool was piloted and refined using a relevant paper identified from the preliminary search (Magee et al., 2012).

### Data Analysis and Presentation

Frequency analysis of publication and study details presented in a table will provide an overview of the scope of evidence pertaining to digital SRHC and GHC for LGBTQI+ youth (aim). Data regarding each of the research questions will be charted on a table and summarised narratively (RQs). For RQ3, where acceptability has been measured qualitatively, data will be extracted verbatim then understood and evaluated using the seven component constructs outlined by Sekhon and colleagues (2017): affective attitude, burden, perceived effectiveness, ethicality, intervention coherence, opportunity costs, and self-efficacy. Where acceptability has been measured quantitatively (e.g., rating of interventions), the relevant numbers will be detailed. Additionally, for RQ4, barriers and facilitators to LGBTQI+ youth accessing and using digital SRHC and GHC will be extracted verbatim and transformed into ‘simple statements’ that capture the essence of the barrier/facilitator.

## Author Contributions

Conceptualisation, JML, CSE, PF, JMD, JG; Methodology, JML, PF, JMD, CSE, JG; Writing - Original Draft, JML; Writing - Review & Editing, JML, JMD, CSE, PF, JG (Brand et al., 2015). All authors have read and agreed to the published version of the manuscript.

## Data Availability

The study is a protocol for a scoping review of publicly available literature.

## Acknowledgements

We would like to acknowledge the contribution of Ron O’Kane (RO), a PhD student at Glasgow Caledonian University (GCU), for being the second reviewer. We would also like to acknowledge Ruth Leiser, a Research Associate at the University of Strathclyde, for peer reviewing this scoping review protocol.

## Appendices

**Appendix A.**
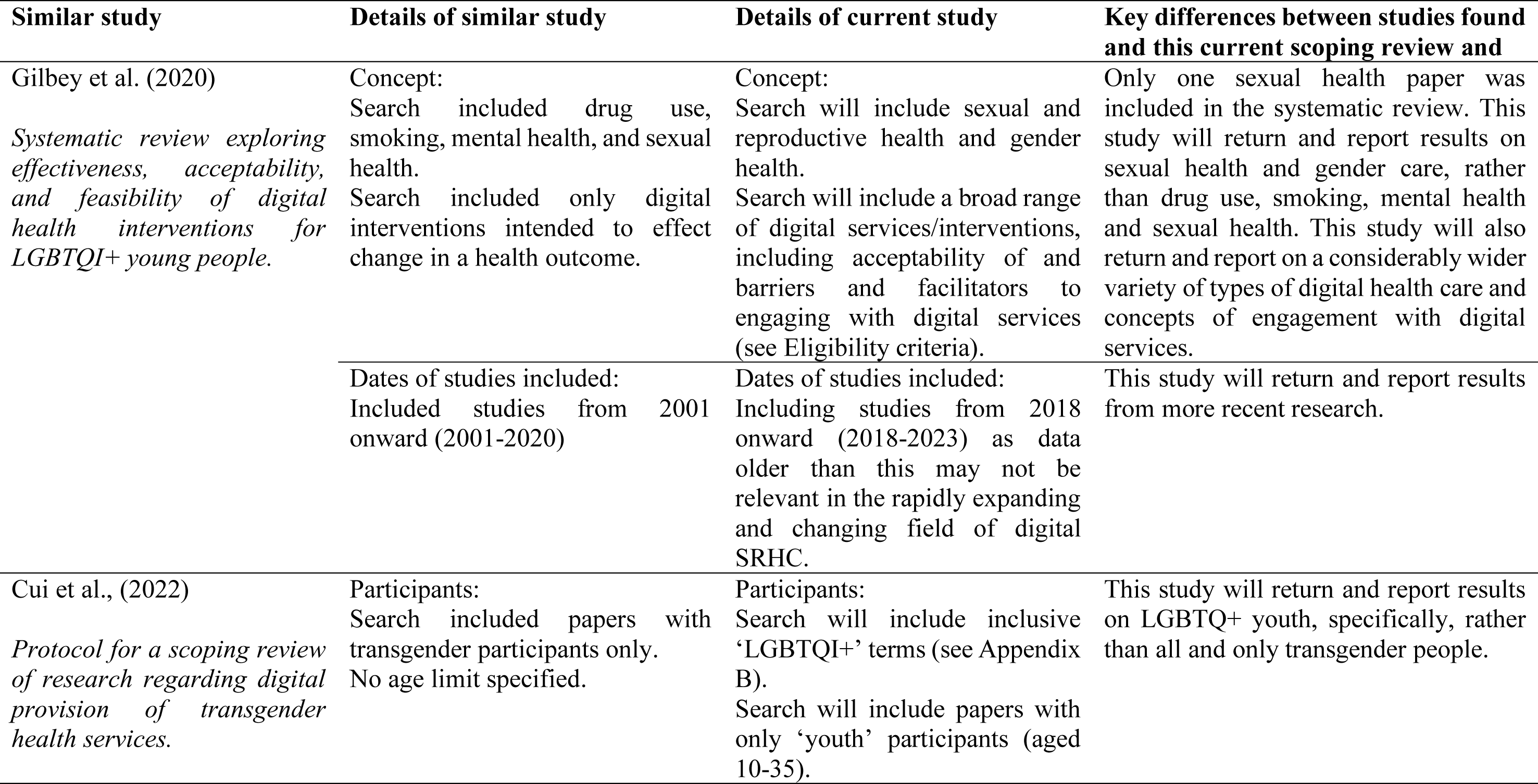
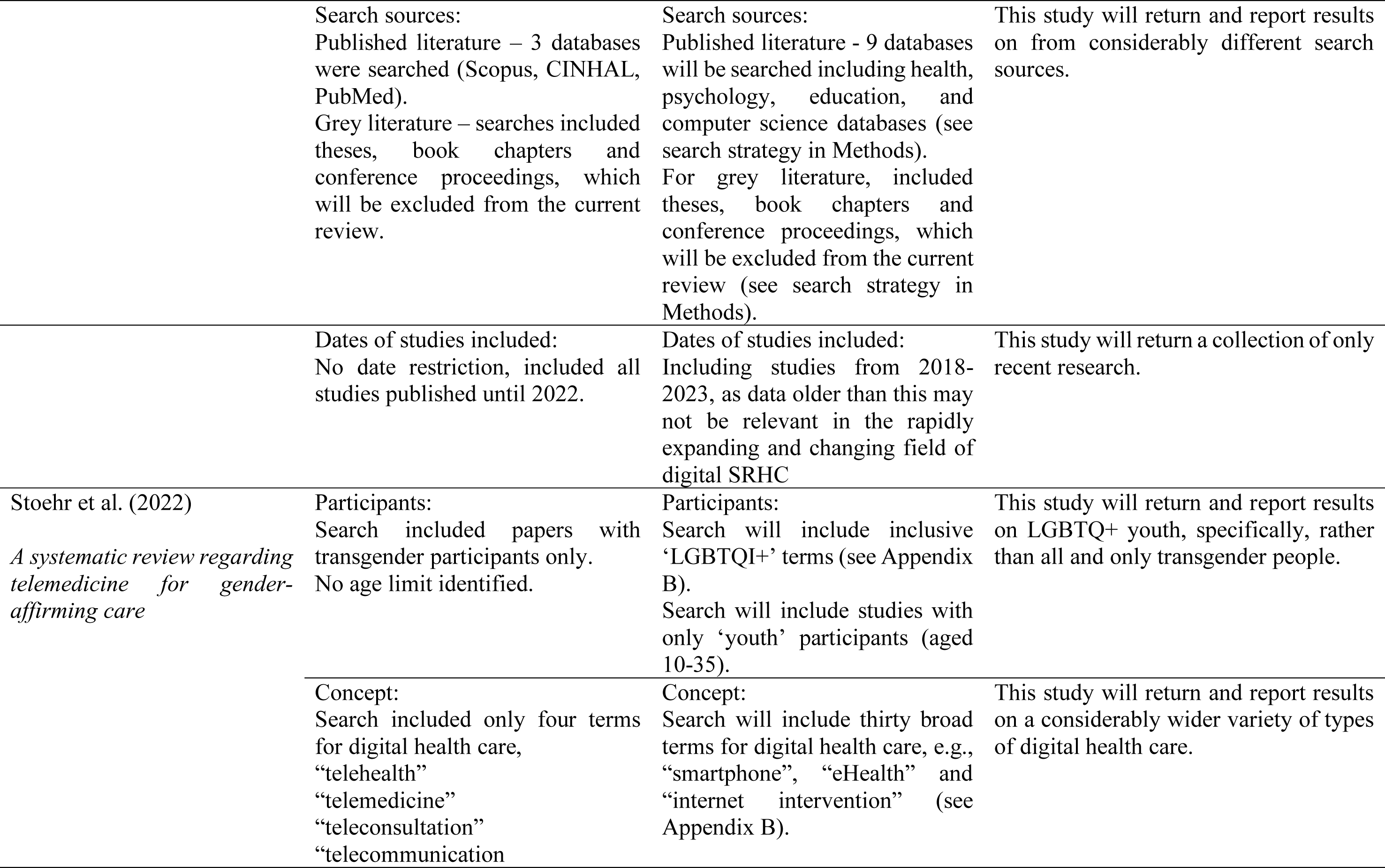
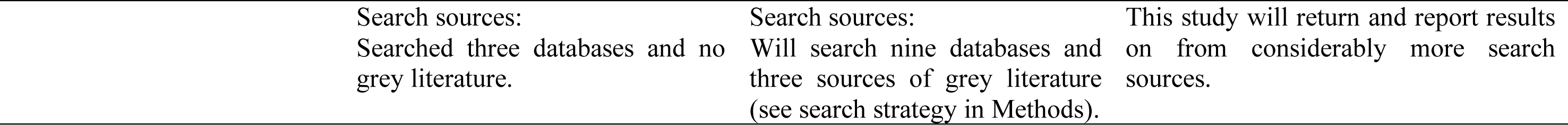
Differences between this current scoping review and similar studies identified.

**Appendix B.**
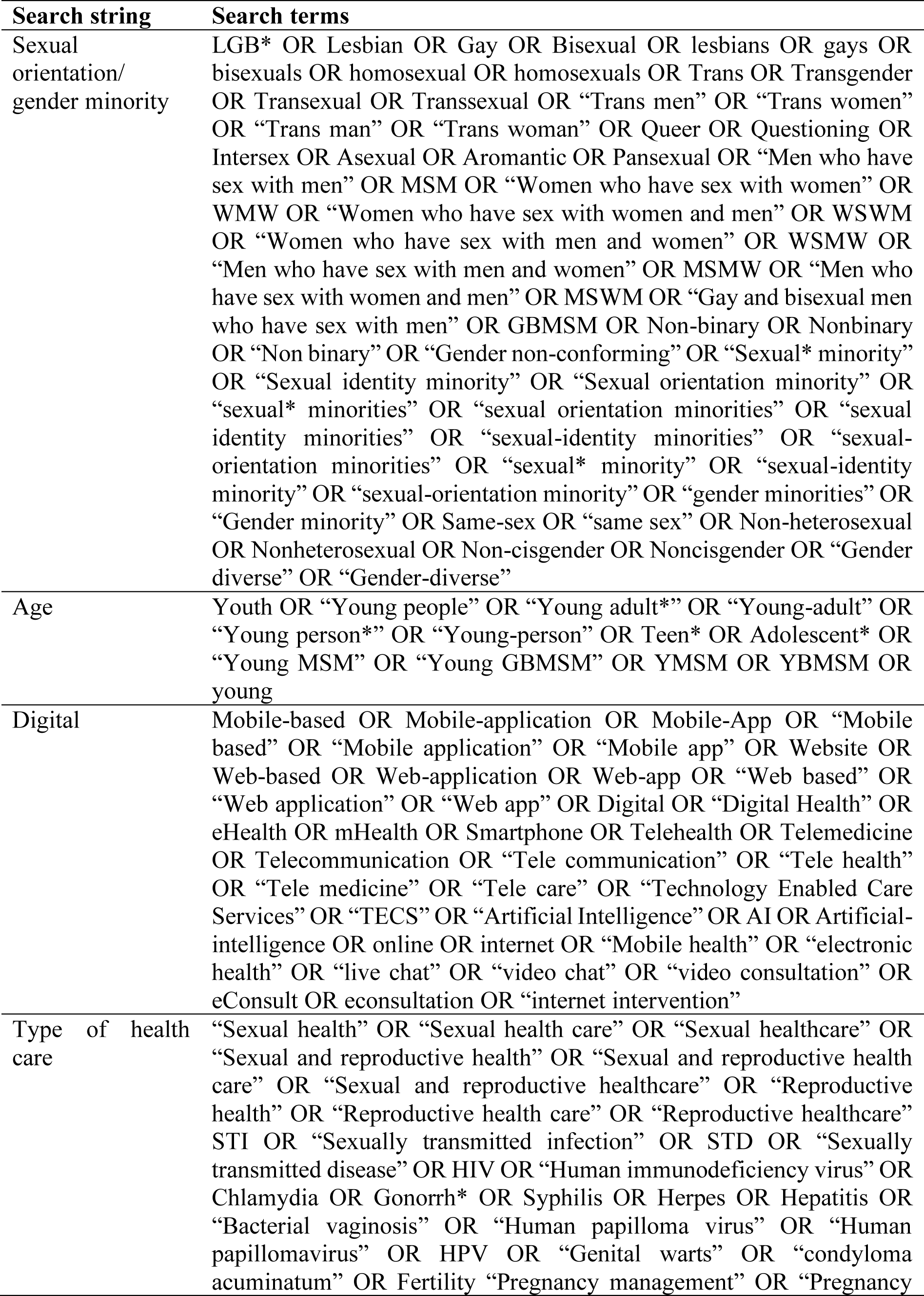
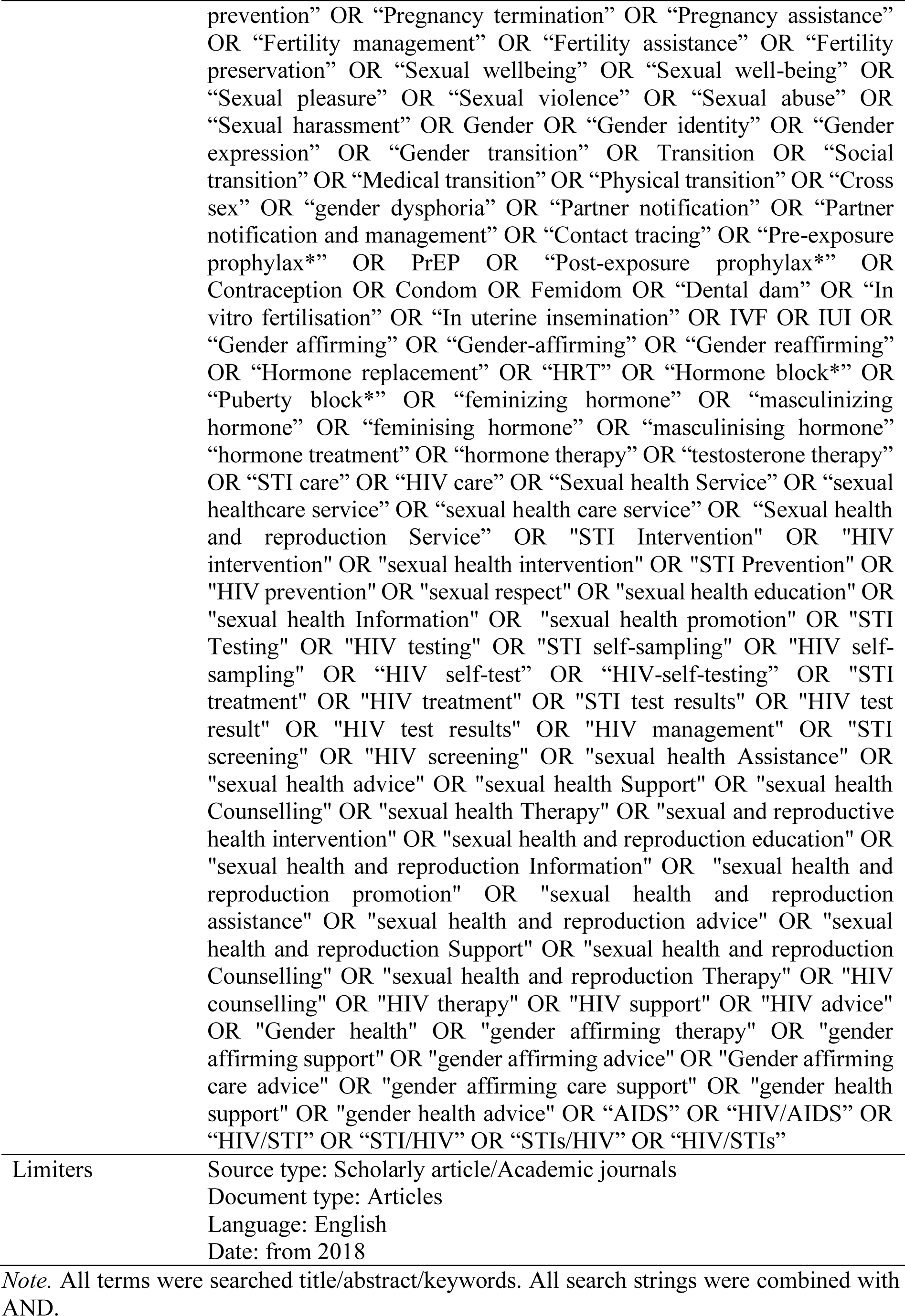
Search terms generated for MEDLINE (EBSCO)

**Appendix C.**
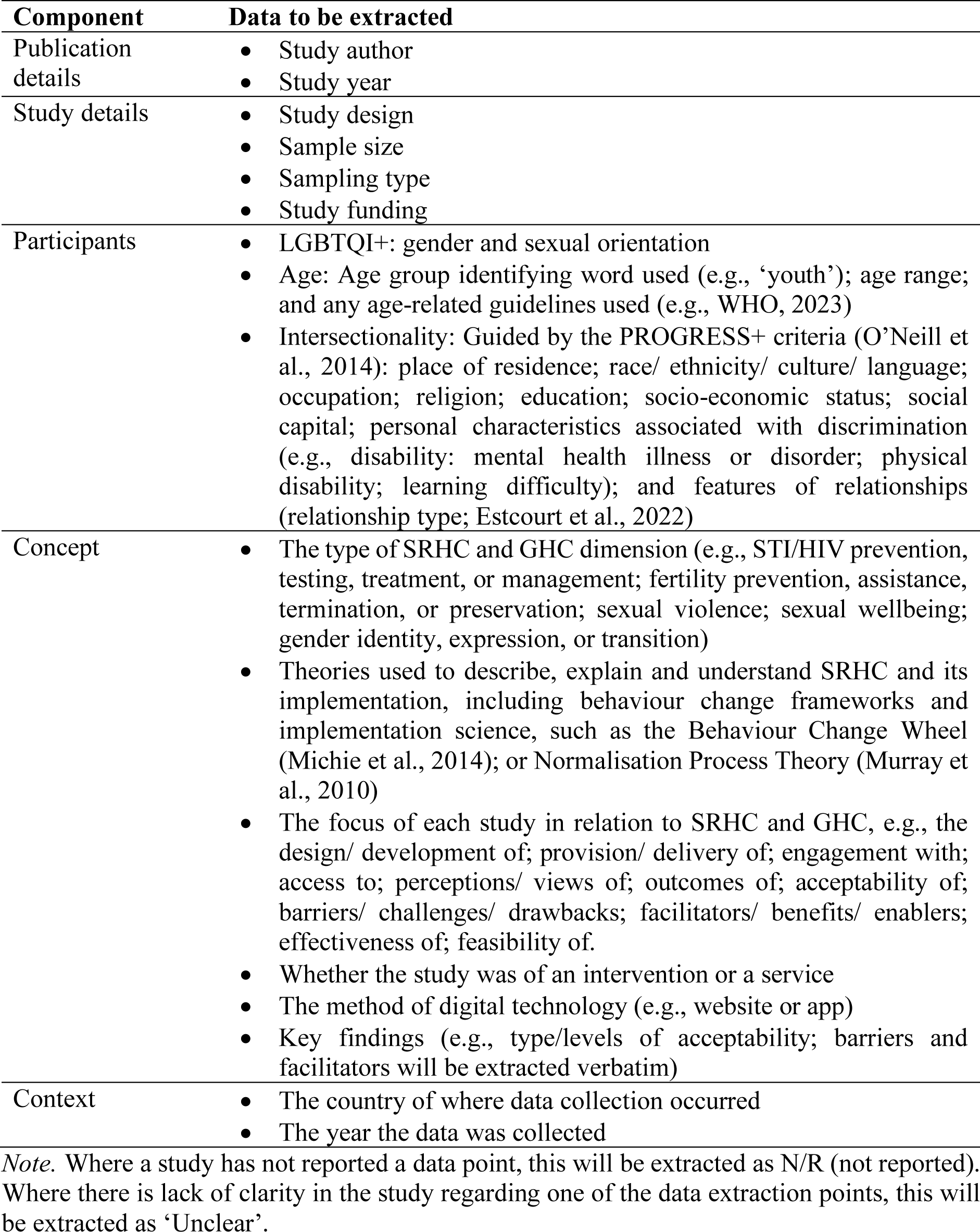
Data to be extracted for scoping review.

| Component | Data to be extracted |
| --- | --- |
| Publication details | • Study author<br>• Study year |
| Study details | • Study design<br>• Sample size<br>• Sampling type<br>• Study funding |
| Participants | • LGBTQI+: gender and sexual orientation<br>• Age: Age group identifying word used (e.g., ‘youth’); age range; and any age-related guidelines used (e.g., WHO, 2023)<br>• Intersectionality: Guided by the PROGRESS+ criteria (O’Neill et al., 2014): place of residence; race/ ethnicity/ culture/ language; occupation; religion; education; socio-economic status; social capital; personal characteristics associated with discrimination (e.g., disability: mental health illness or disorder; physical disability; learning difficulty); and features of relationships (relationship type; Estcourt et al., 2022) |
| Concept | • The type of SRHC and GHC dimension (e.g., STI/HIV prevention, testing, treatment, or management; fertility prevention, assistance, termination, or preservation; sexual violence; sexual wellbeing; gender identity, expression, or transition)<br>• Theories used to describe, explain and understand SRHC and its implementation, including behaviour change frameworks and implementation science, such as the Behaviour Change Wheel (Michie et al., 2014); or Normalisation Process Theory (Murray et al., 2010)<br>• The focus of each study in relation to SRHC and GHC, e.g., the design/ development of; provision/ delivery of; engagement with; access to; perceptions/ views of; outcomes of; acceptability of; barriers/ challenges/ drawbacks; facilitators/ benefits/ enablers; effectiveness of; feasibility of.<br>• Whether the study was of an intervention or a service<br>• The method of digital technology (e.g., website or app)<br>• Key findings (e.g., type/levels of acceptability; barriers and facilitators will be extracted verbatim) |
| Context | • The country of where data collection occurred<br>• The year the data was collected |

*Note.* Where a study has not reported a data point, this will be extracted as N/R (not reported). Where there is lack of clarity in the study regarding one of the data extraction points, this will be extracted as ‘Unclear’.

